# Barriers to Maternal Health-Seeking Behavior in the DRC: The Role of Perceptions, Affordability, and Health System Financing

**DOI:** 10.1101/2025.07.23.25331997

**Authors:** Felipe Montano-Campos, Brittany Hagedorn, Stephen Pope

## Abstract

Efforts to improve maternal health in the Democratic Republic of Congo (DRC) have often focused on expanding access and improving structural readiness. These are critical first steps, but this analysis reveals a critical missing link: women’s perceptions of care—including how they are treated, how much they trust providers, and how affordable care seems—which are more predictive of health-seeking behavior than objective facility infrastructure or actual cost.

Drawing on two rounds of household and facility surveys, we find that care seeking is strongly impacted by perceptions. Women’s perceptions of quality was defined by a composite index that captures interpersonal experiences (e.g., respectful treatment, clear communication, and provider competence), alongside perceptions of facility conditions like cleanliness and drug availability. In regression models, we found that perceived quality was more predictive of care-seeking than structural readiness. Women who perceived care as good were significantly more likely to initiate antenatal care, return for postnatal visits, and seek help for complications—even in under-resourced facilities. These findings highlight the individual-dependent nature of health system interaction: perceived quality modulates behavior even within the same infrastructure.

Similarly, perceptions of affordability better predicted care utilization than objective financial hardship, operating through household coping strategies like asset sales. The type of asset sold mattered: selling food reserves was linked to earlier care but reduced continuity, while selling productive assets enabled sustained engagement. These patterns highlight women’s agency and that affordability is shaped by expectations, vulnerability, and risk assessment.

Health financing models also shaped these dynamics. Performance-based financing (PBF) drove improvements in centrally-incentivized services such as vaccination and laboratory testing, while direct facility financing (DFF) enabled more responsive delivery for high-burden local priorities like malaria care. Crucially, perceptions had a stronger influence on utilization in DFF settings, where facility responsiveness varied more. This suggests that in more adaptive systems, interpersonal trust and perceived quality play a greater role in guiding behavior, highlighting how systems-dependent factors mediate how care is delivered and how patients respond to it.

Improving maternal health in fragile settings requires re-centering perceptions as core indicators of system performance. Respect, clarity, cleanliness, and competence must not only be measured but actively addressed. Financing reforms should empower frontline providers to respond to patient feedback, not just meet centrally imposed targets. And affordability interventions must reflect behavioral realities: what feels affordable is shaped by hardship, expectations, and perceived benefit. Measuring household coping strategies like selling assets can offer insight into the how to mitigate the sacrifices households make to access care and suggest interventions.

Contrary to traditional models of primary healthcare investment, far more attention must be placed on the drivers of perception—friendliness, affordability, trustworthiness—if we aim to expand coverage. Perception data are not mere satisfaction scores; they are proxies for trust, predictors of use, and a missing link in health systems that aspire to be people centered.

## Introduction

Improving maternal health remains a central challenge in low- and middle-income countries (LMICs), where maternal and neonatal morbidity and mortality rates are among the highest globally. Despite substantial investments in health systems, service utilization remains uneven, particularly in fragile contexts like the Democratic Republic of Congo (DRC) [1], [2], [3], [4], [5]. Ensuring that women have access to timely and adequate care during pregnancy, childbirth, and the postnatal period is essential to preventing complications and improving maternal and neonatal outcomes. However, indicators such as late initiation of antenatal care (ANC), failure to complete the recommended four visits (ANC4), low uptake of postnatal care (PNC), and underutilization of services in response to complications remain widespread in the DRC [6], [7], [8].

A growing body of research emphasizes that barriers to maternal health care are not only structural-such as poor infrastructure or long distances-but also behavioral and perceptual [9], [10], [11], [12]. Women may forgo care not because services are unavailable, but because they perceive them as low quality, unaffordable, or unwelcoming [13], [14], [15], [16]. In this context, both perception-based barriers (e.g., concerns about affordability, distrust of providers, fear of mistreatment) and objective facility characteristics (e.g., drug availability, staff presence, equipment) play critical roles in shaping health-seeking behavior [17]. These distinct dimensions may interact but must be empirically disentangled in order to inform targeted interventions. For instance, even well-equipped facilities may be underutilized if patients perceive them as disrespectful or expensive, while positive perceptions may not compensate for actual deficits in readiness or quality.

Disentangling these two sets of barriers – subjective patient perceptions versus objective facility readiness – can contribute to effective policy design, since the required interventions are distinct. From a measurement perspective, separating these effects while also acknowledging their interdependence allows us to understand the degree to which under-utilization is driven by the healthcare environment itself versus how it is experienced and perceived by patients. From a policy standpoint, it informs the optimal investment mix across infrastructure, affordability, trust-building, and patient engagement.

To explore these questions, we opportunistically leverage a dataset from the evaluation of health facility financing models implemented under the DRC’s Health System Strengthening for Better Maternal and Child Health Project (PDSS). The initial study found modest gains in structural quality of health facilities, process quality, and coverage of some health services.

In the PDSS study, there were two financing approaches: Performance-Based Financing (PBF) and unconditional Direct Facility Financing (DFF). These financing models differ not only in how funds are disbursed, but also in the extent to which they introduce performance incentives, monitoring, and management reforms that may affect both structural quality and how services are perceived by patients. In addition to our main analysis examining how perception-based and structural barriers influence maternal health-seeking behavior, we take advantage of variation in financing modality to explore whether these relationships differ by context—providing insight into how supply-side reforms may interact with, mitigate, or exacerbate demand-side barriers.

To conduct this analysis, we use household and facility survey data collected as part of the PDSS impact evaluation. The dataset includes baseline (2015–2016) and endline (2021–2022) surveys from a broad sample of health zones across several provinces. The household surveys include detailed reproductive histories and questions on care-seeking behavior, affordability, and patient experiences among women recently pregnant. These are complemented by health facility assessments that captured data on physical infrastructure, staffing, availability of medicines, service offerings, and quality of care as measured by both provider interviews and clinical observations. The linked structure of the data allows us to trace the relationship between community-level perceptions, facility-level characteristics, and observed utilization of maternal health services in both PBF and DFF settings and over time.

### Conceptual Framework and Theory of Change

Our analysis is guided by a behavioral theory of change that examines how structural factors, household conditions, and individual perceptions jointly shape maternal care-seeking behavior. As shown in Figure 1, we hypothesize that the decision to seek care is not only driven by service availability or true out-of-pocket costs, but also by how women perceive and respond to their environment in the context of social and economic constraints.

**Figure 1.**
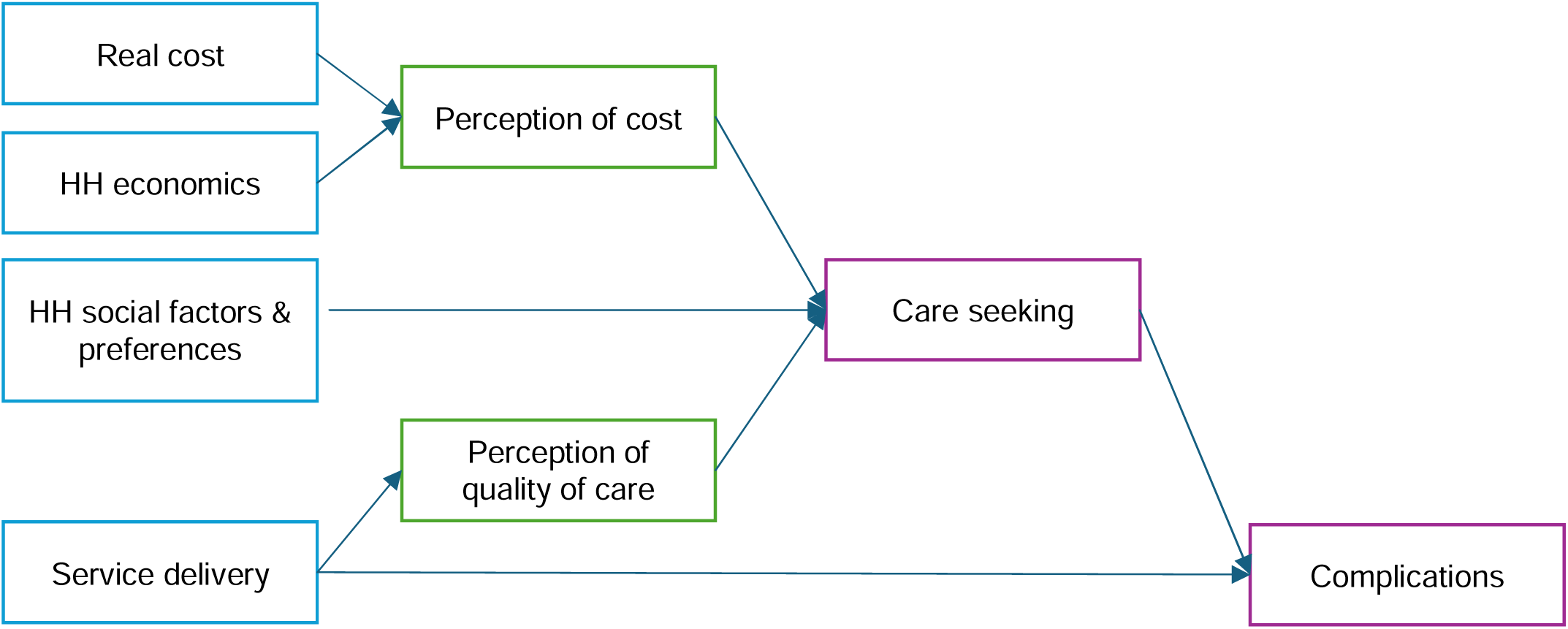
Theory of Change Linking Cost, Quality, and Care-Seeking Pathways.

Per the structure shown above, we conceptualize maternal care-seeking as influenced by three main inputs: (1) household social factors and preferences, (2) perceptions of cost, and (3) perception of quality. These factors affect whether women seek timely and appropriate care during pregnancy, delivery, and the postpartum period.

Each of these dimensions plays a distinct role in shaping behavior and may operate through different mechanisms. Below, we outline how they are defined and how they connect to care-seeking decisions in our analytical framework:

- **Perceptions of care quality** reflect both a) interpersonal experiences such as whether women feel respected, heard, and competently treated and b) assessments of facility conditions, including cleanliness, staff presence, and medicine availability. While these perceptions may be informed by structural realities, they are subjective and socially constructed as well as being time-delayed (i.e., previous experiences influence current perceptions).
- **Perceptions of affordability** are shaped not only by real service and medicine costs but also by household-level financial strain. Women who face high out-of-pocket expenses or who must borrow money or sell assets to afford care may be more likely to perceive costs as prohibitive, regardless of the nominal fee structure.
- **Household social factors and preferences**, including an individual’s trust in traditional medicine directly influence demand for services and may affect how women perceive both cost and quality.

These dimensions interact to influence care-seeking behavior, determining whether, when, and how women engage with the health system. Understanding how each barrier operates, and where perceptions align or diverge from reality, helps to identify which constraints are addressable and through what mechanisms. This in turn can be used to design policy changes and optimize investments in the healthcare system.

Finally, maternal complications are conceptualized as a downstream outcome of this system. Complications are shaped by both the timeliness and completeness of care-seeking, as well as the quality of services received once care is accessed. They thus reflect the cumulative influence of both demand-side barriers and supply-side readiness.

### Data

#### Survey design and implementation

This study relies on two waves of primary data collection carried out in the DRC as part of a large-scale effort to evaluate health system performance and service delivery outcomes. The data include detailed household and facility-level surveys conducted at baseline (2015–2016) and endline (2021–2022), designed to capture both the demand- and supply-side conditions shaping maternal and child health care.

The baseline survey was conducted between June 2015 and March 2016 across 133 health zones in 17 provincial health districts (DPS). It included data from over 6,500 randomly selected households, targeting women aged 15 to 49 who had been pregnant in the preceding two years. These household interviews collected information on reproductive histories, care-seeking behavior, experiences with health services, household assets and health expenditures, and perceptions of care affordability and quality. In each health zone, surveys were also administered in five randomly selected health centers and a sample of general referral hospitals, yielding comprehensive data on facility infrastructure, staffing, equipment, drug availability, and service readiness. Additional modules included provider interviews and technical knowledge assessments using clinical vignettes.

The endline survey, conducted in 2021–2022, followed a comparable design but focused on a subset of 58 health zones in six provinces that had been part of a randomized impact evaluation. Household data again focused on recently pregnant women, allowing for detailed tracking of maternal health behaviors over time. Facility assessments mirrored those of the baseline and captured structural readiness, quality metrics, staffing, and availability of maternal and newborn services. This longitudinal design, with linked household and facility surveys, enables direct measurement of changes in perceived and objective barriers to care. Women were linked to the closest health facility to their household, which served as a proxy for their usual source of care. Since fewer than 11% of respondents reported using a different facility than the nearest one, this approach provides a reasonable approximation of where care was actually received.

This dataset originates from a rigorous impact evaluation of PBF implemented under the Health System Strengthening for Better Maternal and Child Health Results Project (PDSS). Introduced in late 2016, the PBF reform tied facility payments to verified performance on service delivery and quality indicators. A cluster-randomized trial was embedded in the PDSS design, with health zones randomly assigned to either PBF or to a control group receiving equivalent unconditional transfers-referred to as direct facility financing (DFF). Both groups of facilities received the same average funding, though there were facility-level adjustments to account for equity and rurality with the intention to compensate for the real costs of delivering healthcare in the DRC, where logistics are expensive and passable roads can be scarce. This mechanism meant that facilities with catchment populations that were under-served and remote received more funding than those that were not, regardless of study arm.

While the data were generated in the context of evaluating this financing reform, the focus of our study is to understand how perceived affordability, perceived quality of care, and household preferences shape maternal health-seeking behavior. We use the data to examine how these barriers relate to outcomes such as antenatal care initiation and completion, postnatal care, and care for pregnancy-related complications. Importantly, the PBF and DFF rollout provides us with a natural opportunity to assess whether and how these barriers evolved differently over time and across financing models. By comparing health zones before and after the introduction of financing reforms and across facilities operating under different incentive structures, we are able to explore how both perception-based and structural barriers affect behaviors in different health system environments.

#### Outcomes of Interest

We examine a range of maternal health outcomes that capture different aspects of care-seeking behavior, including service utilization, timeliness of access, and engagement with the health system around complications. We retain the individual-level data for analysis, so all outcomes are binary indicators (e.g., received ANC or not) based on self-reported responses from recently pregnant women interviewed during the household surveys.

**Care utilization** is captured through three key indicators:

- Receipt of any antenatal care (ANC1), defined as attending at least one ANC visit during the most recent pregnancy;
- Adequate antenatal care (ANC4), defined as completing four or more ANC visits; and
- Receipt of any postnatal care (PNC), defined as attending a postnatal visit within two months of delivery.

**Timeliness of care** is assessed based on the World Health Organization’s (WHO) recommendation for early initiation and timely follow-up care, defined as:

- Whether the first ANC visit occurred six or more months into pregnancy (late ANC), and
- Whether postnatal care was received more than seven days after delivery (late PNC).

We examine outcomes related to **pregnancy complications** that reflect both preventive engagement with the health system and reactive care-seeking when complications arise. These include:

- Whether the respondent reported discussing potential complications during prenatal care,
- Whether she received advice on where to go for care in the event of a complication, and
- Whether she experienced any complications during pregnancy or delivery.

#### Key Explanatory Variables

##### Quality of Care Perception Index

To capture women’s perceptions of quality of care, we constructed a composite index using a set of survey questions asked to recently pregnant women about their experiences with the health system. These items focused on how women subjectively evaluate the care they received and the environment in which it was provided. Rather than analyzing each item individually, we applied principal component analysis (PCA) to create a summary score representing overall perceived quality. The perception variables used to generate the single index reflect both interpersonal aspects and structural dimensions of care. These include:

###### Interpersonal experience with providers

The survey asked women whether they felt well received at the health facility, whether they were treated with respect, whether providers explained things clearly, whether they had the opportunity to ask questions, and whether they felt the staff were competent. These items reflect women’s perceptions of patient–provider interaction, including communication quality, attentiveness, and emotional support.

###### Perceptions of facility conditions and capacity

Respondents were also asked about the physical and service environment of the facility. This included whether the facility was clean, had essential medicines and functional equipment, offered diagnostic services, provided effective treatments, and was open at convenient hours. These indicators reflect the woman’s subjective evaluation of the facility’s structural readiness to deliver quality care.

After generating a continuous score from the PCA and normalizing it from 0 to 1 (See Figures 2, below), we categorized the index into three levels (poor, fair, and good) to make interpretation easier. This classification was based on the distribution of the PCA scores in the sample, where values below the 25th percentile were classified as “poor,” those between the 25th and 75th percentiles as “fair,” and those above the 75th percentile as “good.” We conducted a validation exercise to assess the internal consistency and distributional properties of the index, as detailed in Appendix A. To ensure that the index appropriately reflects underlying variation in perception items, we conducted a validation exercise using the baseline household survey data. For each individual perception variable, we grouped women by their reported rating—poor, fair, or good—and then plotted the average value of the overall perceived quality index across these three levels. As shown in Figure A1, higher ratings on each individual item are associated with higher values of the composite index, confirming that the index aligns consistently with the rating structure across all perception dimensions. Variables with steeper gradients across rating levels contribute more heavily to the first principal component and thus have greater influence in shaping the overall perception score.

**Figure 2.**
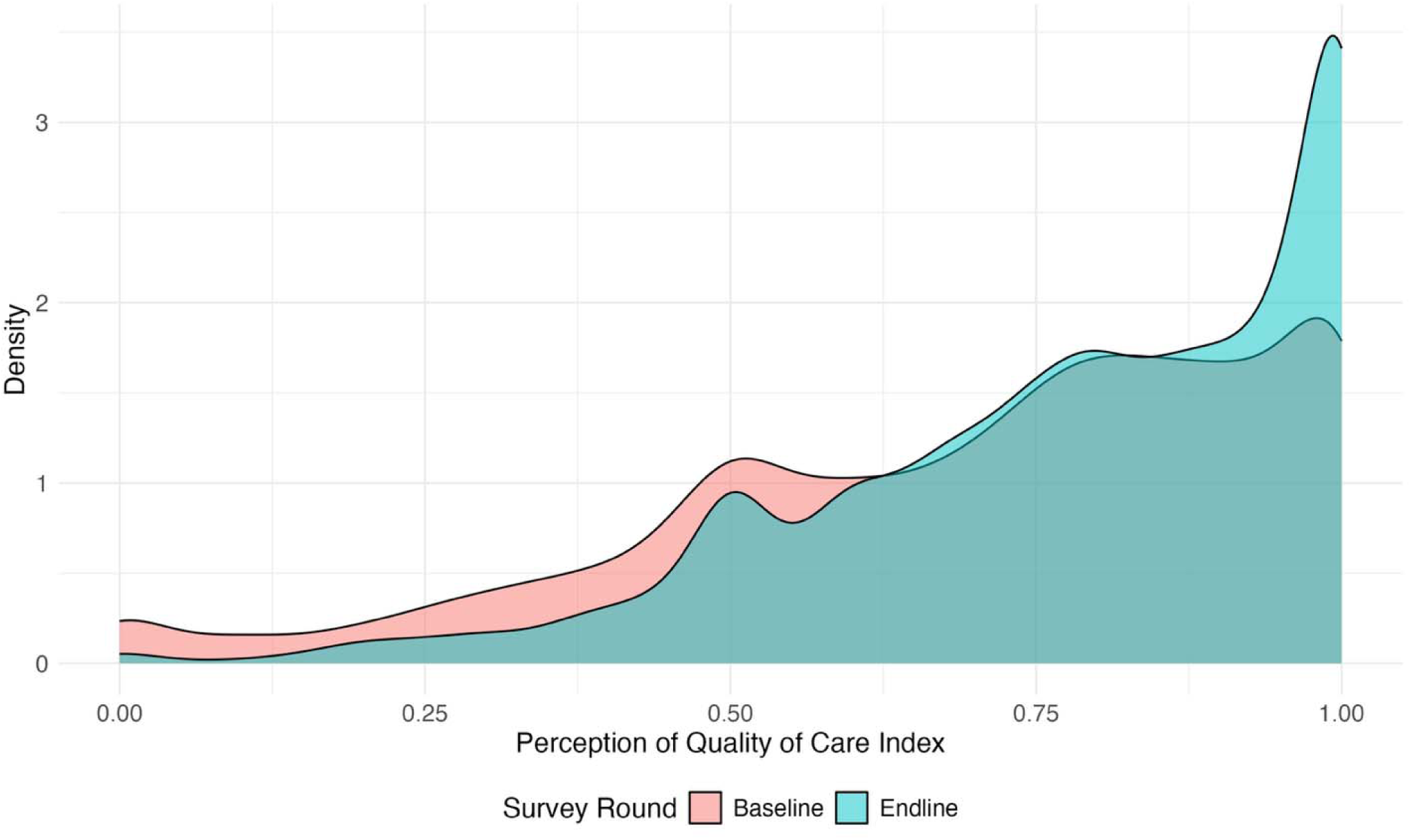
Distribution of Perceived Quality of Care Index Scores at Baseline and Endline. *Density plot of normalized index (0–1 scale), stratified by survey round*.

This index was constructed using data from both the baseline and endline household surveys.

##### Facility Readiness Indices

To capture objective supply-side variation in facility capacity, we constructed a set of indices that summarize facility readiness using data from the health facility surveys. Each index is based on a set of variables capturing key characteristics of a facility relevant to a specific service domain. For each domain, we constructed a single composite index using PCA to summarize how well a facility performs across all related indicators.

We created separate indices for the following domains, aligned with the facility survey structure: Laboratory Test Availability, Vaccination & Cold Chain Services, Prenatal Services, Childbirth & Postpartum Services, Tuberculosis Services, Malaria Services, Available Equipment, and Essential Medicines Availability. Each index was normalized to range from 0 to 1, with higher values indicating better structural readiness in the corresponding domain.

For ease of interpretation and policy relevance, we further categorized each continuous index into a binary indicator identifying facilities that were high performing in that domain. Facilities whose index values fell within the top third of the distribution were classified as high-performing and assigned a value of one, while all others were coded as zero (Sensitivity analyses using alternative thresholds—such as the top quarter or top half—yielded similar patterns of association). This binary classification provides a clear and interpretable measure of which facilities stood out in terms of service-specific readiness and allows us to assess how structural quality may influence care-seeking behavior. Additional details on the variables included in each index are provided in Appendix B.

These indices were constructed using data from both the baseline and endline facility surveys, which incorporated direct facility assessments and provider interviews.

## Methods and Empirical Approach

This section outlines the empirical strategies used to examine how different barriers-perceived quality, structural readiness, and financial hardship-affect maternal care-seeking. We begin with a general modeling framework used across multiple outcome analyses. In the subsections that follow, we outline specific analytic approaches tailored to the relationships between structural and perceived quality (Section 4.2) and between financial hardship, affordability, and antenatal care-seeking (Section 4.3).

### Modeling Framework

All outcomes in our analysis are binary indicators of maternal health behavior or experience, such as receipt of antenatal care, delayed initiation of care, postnatal care utilization, or engagement with providers around pregnancy complications. To estimate the association between key behavioral or structural barriers and these outcomes, we use logistic regression models of the following form:

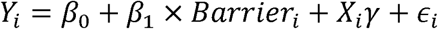

where *Y_i_* is the binary outcome for woman *i*, and *Barrier_ij_* represents our primary variable of interest, which may include perceived quality of care across different domains, perceived affordability, financial hardship, or other behavioral constraints. The matrix *X_ij_* includes a rich set of sociodemographic controls: woman’s age, pregnancy status, marital status, education level, religion, household wealth (quartile), and geographic location captured by province fixed effects. The coefficient of interest, *β*_1_, captures the association between each barrier and the likelihood of engaging in a given maternal health behavior, conditional on sociodemographic characteristics and geographic context.

This general framework is used to analyze multiple maternal health-seeking outcomes previously described, including antenatal care (ANC1 and ANC4), postnatal care, timeliness, and engagement with providers around complications. The explanatory variables in this set of models include individual perception variables (e.g., cleanliness, respect, opportunity to ask questions) and household-level preference indicators (e.g., preference for traditional medicine). Results from this analysis are presented in Section 5.2.

While Section 4.1 presents the general modeling approach used to explore how individual-level perceptions and preferences relate to maternal health behaviors, the following sections build on this foundation by examining three key drivers of behavioral and structural outcomes:

- **Women’s perceptions** (4.2)

◦ How closely linked are women’s *perceptions* of quality and *objective indicators* of facility readiness?
◦ Do perceptions drive health seeking, independent of structural quality?
- **Financing models** (4.3)

◦ Supply: How does financing impact facility readiness to provide care?
◦ Demand: How does financing affect perceived quality and care-seeking?
- **Affordability & Agency** (4.4)

◦ How do financial hardship and affordability drive antenatal care-seeking?
◦ How do coping strategies, such as selling assets, influence decisions of how to engage with the health system?
◦ This section highlights the role of agency and economic decision-making under realistic household budget constraints.

### Linking Perceived Quality, Structural Quality, and Care-Seeking Behavior

We first test whether perceived quality of care is meaningfully linked to the actual structural conditions of the health facilities women use. If perceptions were completely unrelated to observable facility readiness, it would be difficult to interpret the influence of perceptions on health-seeking behavior or to identify actionable policy responses. To explore this, we examine whether women’s ratings of care are systematically associated with objective indicators of service availability and infrastructure. At the same time, we recognize that perceptions are not purely determined by structural conditions—they are also shaped by interpersonal experiences, expectations, and trust in the health system. Our goal is therefore not to equate perception with readiness, but to establish the degree to which there is a measurable connection between them. To do so, we estimate the following model:

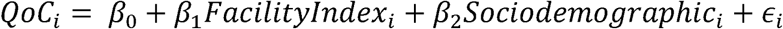

Where *QoC_i_* represents the perceived quality index reported by individual woman *i*, *Facilitylndex_i_* includes readiness scores across key domains such as laboratory services, vaccination and cold chain, prenatal and delivery care, tuberculosis and malaria services, and operational equipment, and *Sociodemographic_i_* includes fixed effects for the woman’s age, marital status, household wealth (quartile), and province. This model allows us to examine the degree to which women’s perceptions of care reflect the objective measures of structural quality.

Once we understand the relationship between perception and structural quality, we next estimate the relationship between perceived quality of care and maternal health-seeking outcomes, including use of antenatal care (ANC1, ANC4), postnatal care, timeliness of care, and care-seeking for complications. In these models, we include both perceived quality and facility readiness indicators to separately identify their associations with behavior. The model is specified as:

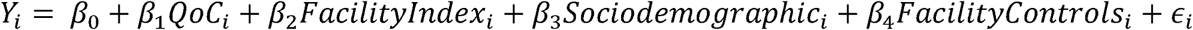

Where *Y_i_* is a binary indicator for maternal care-seeking behavior, *QoC_i_* is the categorized perceived quality index, *Facilitylndex_i_* includes the relevant domain-specific readiness scores, and *FacilityControls_i_* includes broader facility-level characteristics such as infrastructure investment, ownership, and funding source. All models include the same individual-level controls and province fixed effects. By including both perceived quality and objective structural quality (i.e., facility readiness) in the model, we are able to assess their independent effects. This distinction is important: structural investments may not translate into increased care-seeking if patients still perceive care as poor, while strong perceptions of quality may drive utilization even when resources are limited. Disentangling these two dimensions helps inform whether policy should prioritize improving facility conditions, building trust and communication, or both.

### Assessing the Impact of Financing Models on Facility Improvements

To evaluate how financing mechanisms influenced facility performance over time, we employed a difference-in-differences (DiD) approach comparing changes between baseline and endline among facilities assigned to either PBF or DFF. This analysis was restricted to the 269 facilities observed in both survey waves to ensure a consistent sample.

We used normalized, PCA-based indices of service readiness and availability (see Section 3.3) to summarize facility characteristics across key domains, including prenatal care, laboratory testing, vaccination, tuberculosis services, and malaria services. For perceived quality of care, we calculated the average score at the facility level by aggregating responses from all women who reported that facility as their primary source of maternal care, enabling alignment between household perceptions and facility-level performance measures.

We estimated DiD models using facility fixed effects and clustered standard errors to account for time-invariant unobserved heterogeneity. The model specification is as follows:

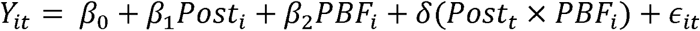

where *Y_it_* denotes the outcome of interest for facility *i* at time *t*; *Post_t_* is an indicator for the endline period; and *PBF_i_* is a binary indicator equal to 1 for facilities assigned to the PBF arm (0 for DFF). The interaction term *Post_t_ X PBF_i_* captures the difference-in-differences estimate. The coefficient of interest, *δ*, measures the relative change in outcomes for PBF facilities compared to DFF facilities between baseline and endline. A positive *δ* indicates that PBF facilities improved more—or declined less—than DFF facilities, while a negative value indicates greater relative gains under DFF.

The estimated effects are expressed as a percentage of the baseline mean for each outcome, allowing for comparison across indicators.

### Linking Financial Hardship, Perceived Affordability, and Antenatal Care-Seeking Behavior

We next examine the role of financial barriers in influencing maternal health-seeking behavior, with a specific focus on antenatal care (ANC). The objective is to disentangle whether financial hardship influences care-seeking directly by constraining a household’s ability to act or indirectly through perceived affordability, defined as a woman’s subjective evaluation of whether the cost of services and medicines is acceptable. This approach allows us to assess the mechanisms through which financial constraints shape healthcare decisions.

We begin by estimating the association between financial hardship and perceptions of affordability. Perceived affordability was measured in the household survey using a three-level categorical variable that captures whether the woman rated costs as unacceptable, average, or acceptable. To account for the ordinal nature of this outcome, we estimate an ordered logistic regression model of the form:

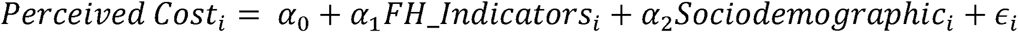

Where *Perceived Cost_i_* is the perceived affordability rating for woman *i*, and *FH_lndicators_i_* includes binary indicators for that woman’s household financial hardship experiences, such as selling assets, borrowing money, or receiving gifts to pay for care. In this model, *α*_1_ reflects the direct effect of how financial hardship affects perception of cost/affordability.

We then estimate the direct association between financial hardship and perceived cost/affordability with the likelihood of receiving any antenatal care (ANC1). This is captured in the following specification:

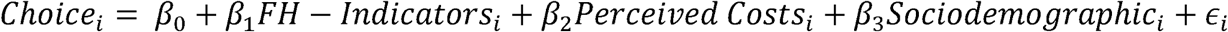

In this model, the coefficient *β*_1_ reflects the direct effect of financial hardship on care-seeking, while *β*_2_ captures the direct effect of perceived cost/affordability on care-seeking.

To quantify the mechanisms through which financial hardship affects care-seeking, we apply a mediation framework. Specifically, we calculate the indirect effect of financial hardship on ANC use—operating through perceptions of affordability—as the product of *α*_1_ × *β*_2_, where α comes from the first regression and *β*_2_ from the second. The total effect of financial hardship is the sum of the direct pathway (*β*_1_) and the indirect pathway (*α*_1_ × *β*_2_).

## Results

### Study Sample and Descriptive Statistics

The analysis draws on two waves of household survey data collected as part of the PDSS evaluation. The baseline survey (2015–2016) includes 6,744 women from 6,556 households across 133 health zones and 17 provinces (Provincial Health Divisions). The endline survey (2021–2022) includes 10,141 women from 8,510 households, sampled in 58 health zones across 6 provinces. All respondents were women who were recently pregnant or gave birth in the two years preceding the survey.

To complement the household data, we also use facility survey data to capture structural readiness. The baseline survey includes 756 health facilities, while the endline covers 346 facilities, distributed across the same study areas as the household survey.

To provide context for the main analyses, we present key demographic and socioeconomic characteristics from the baseline sample. Most women were married (57% in monogamous unions and 11% in polygamous unions), with an additional 21% living with a partner (Figure 3). The sample is broadly distributed across reproductive ages, with the largest shares between 20 and 30 years old (Figure 3). Educational attainment is low: 32% of women had no formal education, and only 1% had reached higher education (Figure 3). The wealth distribution is heavily skewed, with a large proportion of households clustered in the lowest end of the wealth index (Figure 3).

**Figure 3.**
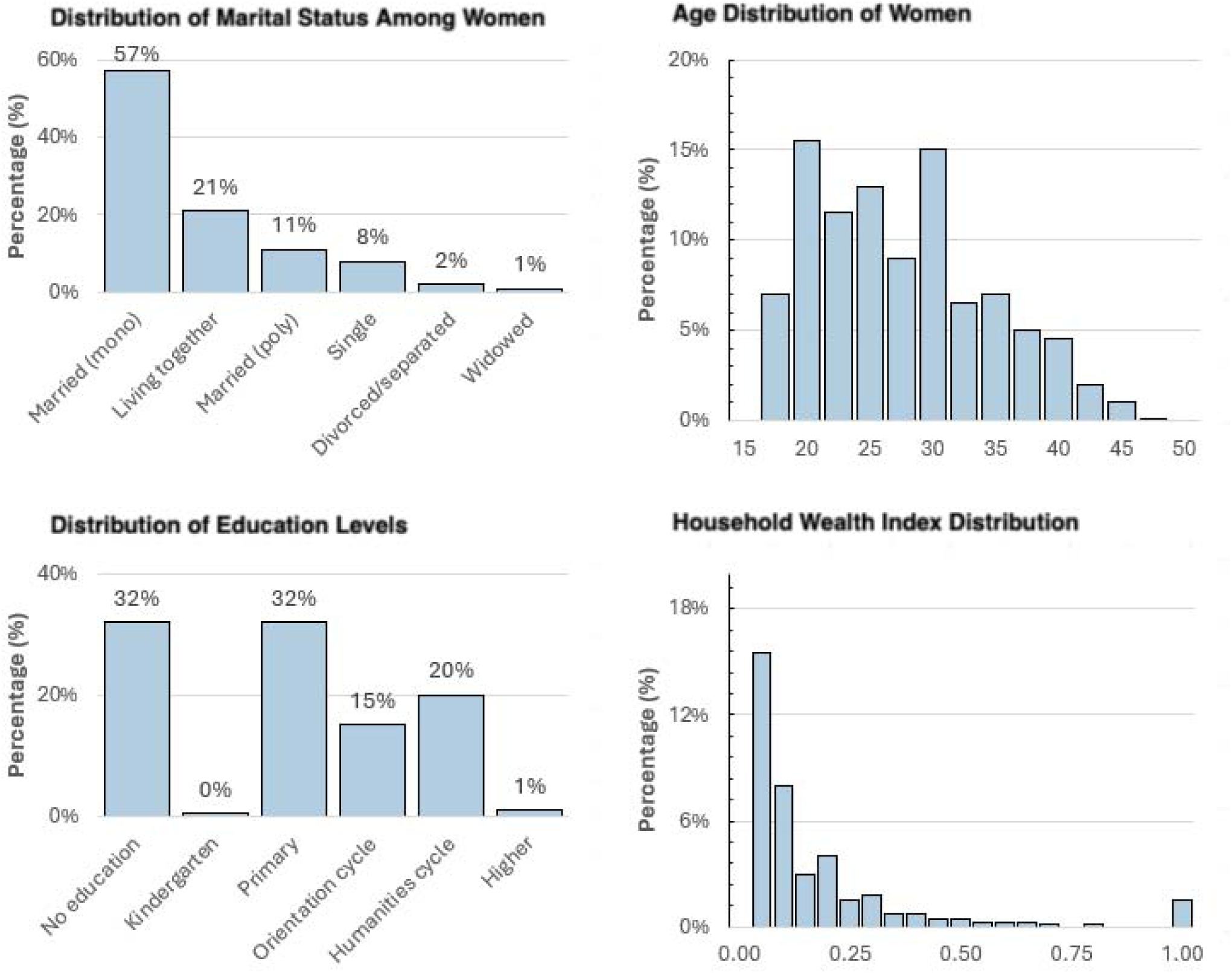
Baseline Characteristics of Women (Baseline Sample)

We now turn to the main findings, beginning with the relationship between perceived quality of care and maternal health-seeking outcomes.

### The Role of Perceptions and Preferences in Maternal Care-Seeking

This section presents findings from the baseline survey on cultural preferences and individual perception variables, based on direct survey responses. These measures capture beliefs and patient-reported experiences-such as trust in traditional medicine or perceptions of cleanliness and communication-that may influence maternal care-seeking behavior. This analysis corresponds to the general modeling framework described in Section 4.1, where binary maternal health outcomes are regressed on individual-level perception and preference variables Importantly, these results reflect individual indicators only; findings related to the composite perceived quality of care index are presented separately in Section 5.3.

Preference for traditional medicine was significantly associated with reduced care-seeking across multiple outcomes. Compared to those who did not report a preference for traditional medicine, women who did were:

- 75% less likely to seek antenatal care (ANC1) (OR = 0.25; 95% CI: 0.08–0.80; p = 0.019)
- 62% less likely to discuss complications during an ANC visit (OR = 0.38; 95% CI: 0.15– 0.93; p = 0.034)
- 70% less likely to receive postnatal care (OR = 0.30; 95% CI: 0.09–0.92; p = 0.035)
- 58% less likely to seek care within 24 hours of experiencing a complication (OR = 0.42; 95% CI: 0.21–0.83; p = 0.013).

These associations were adjusted for age, marital status, education, religion, wealth, and geographic location.

Several specific perception variables were also associated with improved care-seeking and outcomes. Women who perceived facilities as clean were more than twice as likely to seek care for complications (OR = 2.08; 95% CI: 1.04–4.15), and those who reported positive impressions of examination equipment were 146% more likely to do so (OR = 2.46; 95% CI: 1.28–4.74).

Perception of the opportunity to ask questions during care was associated with a 28% lower likelihood of reporting complications during pregnancy (OR = 0.72; 95% CI: 0.52–0.98).

Additionally, women who perceived the distance to a facility as acceptable were 20% more likely to initiate antenatal care earlier in pregnancy (OR = 0.80; 95% CI: 0.69–0.94).

A full set of results across all perception domains and maternal health outcomes is presented in Table 1. Overall, perceptions were most consistently associated with care-seeking behaviors for ANC1 and PNC, where nearly all perception variables showed significant positive associations. In contrast, only few perceptions were significantly linked to care completion (ANC4) or timeliness (late ANC), suggesting that perceptions may be more influential in shaping whether women engage with the health system at all, rather than how early or consistently they do so.

**Table 1.** Adjusted Odds Ratios (ORs) and 95% Confidence Intervals for the Association Between Individual Perception Variables and Maternal Health-Seeking Outcomes. *Coefficients estimate the effect of a patient having a positive “acceptable” perception on care utilization, relative to “not acceptable” perception. All models are adjusted for age, pregnancy status, marital status, education, religion, household wealth, and province fixed effects. Green cells indicate statistically significant results at p < 0.01; light green indicates p < 0.05. ANC = antenatal care. PNC = postnatal care. 1=first visit. 4=at least four visits. Late = first visit initiated at six months or later*.

| Perception | Care Seeking |  | Timeliness | Completion |
| --- | --- | --- | --- | --- |
| Outcome: | ANC1 | PNC | Late ANC1 | ANC4 |
| Med Availability | 1.61 (1.31, 1.98) | 1.24 (0.96, 1.60) | 0.88 (0.74, 1.04) | 0.81 (0.69, 0.95) |
| Opening Hours | 1.56 (1.18, 2.03) | 2.84 (1.72, 5.01) | 0.84 (0.66, 1.09) | 1.18 (0.92, 1.50) |
| Patient Reception | 1.54 (1.17, 2.02) | 1.76 (1.10, 2.97) | 0.75 (0.58, 0.98) | 0.92 (0.71, 1.19) |
| Treat Effectiveness | 1.49 (1.20, 1.85) | 3.16 (2.07, 5.02) | 1.00 (0.82, 1.23) | 1.05 (0.86, 1.29) |
| Medicine Cost | 1.43 (1.11, 1.86) | 1.90 (1.44, 2.49) | 0.85 (0.70, 1.03) | 1.03 (0.85, 1.24) |
| Respect Patients | 1.42 (1.10, 1.83) | 2.47 (1.52, 4.29) | 0.94 (0.74, 1.19) | 0.87 (0.68, 1.09) |
| Cleanliness | 1.39 (1.14, 1.69) | 1.01 (0.76, 1.35) | 1.01 (0.85, 1.21) | 0.75 (0.63, 0.89) |
| Service Cost | 1.37 (1.11, 1.70) | 1.63 (1.26, 2.12) | 0.86 (0.72, 1.02) | 0.98 (0.83, 1.16) |
| Ask Questions | 1.36 (1.07, 1.71) | 1.92 (1.25, 3.06) | 0.88 (0.71, 1.10) | 0.84 (0.67, 1.04) |
| Interaction re: Illness | 1.31 (1.03, 1.66) | 2.08 (1.34, 3.39) | 0.97 (0.77, 1.22) | 0.76 (0.60, 0.95) |
| Exam Equipment | 1.29 (1.06, 1.57) | 1.31 (1.00, 1.73) | 0.91 (0.76, 1.07) | 0.73 (0.62, 0.86) |
| Staff Competence | 1.20 (0.92, 1.57) | 2.19 (1.28, 4.04) | 0.80 (0.63, 1.03) | 1.01 (0.79, 1.28) |
| Diagnostic Methods | 1.09 (0.88, 1.35) | 1.53 (1.10, 2.17) | 0.92 (0.76, 1.10) | 0.92 (0.78, 1.08) |

### Structural Readiness and Its Influence on Perceived Quality of Care

As outlined in Section 4.2, we assess whether structural facility readiness is associated with women’s perceptions of care quality. Using baseline data, we test whether higher objective readiness across service domains predicts better perceived quality (categorized as “poor,” “fair,” or “good”), controlling for sociodemographic factors and province fixed effects.

The results show that several domains of structural readiness are positively associated with patient perception. Specifically, patients that attended facilities in the top tertile for operational equipment were 40% more likely to rate overall perceived quality as “good” (OR = 1.40; 95% CI: 1.25–1.58). Similar positive associations were observed for malaria services (OR = 1.23; 95% CI: 1.10–1.38), laboratory testing (OR = 1.30; 95% CI: 1.15–1.47), and tuberculosis services (OR = 1.17; 95% CI: 1.04–1.31). The associations for prenatal, childbirth, and vaccination service domains were smaller and not statistically significant at p<0.05. These findings suggest that supply-side investments in diagnostics, equipment, and malaria service readiness may be more salient to patient perceptions in this context than other service inputs.

### Impact of Perceived Quality of Care on Health-Seeking Behavior

We now examine the association between perceived quality of care and key maternal health behaviors, using baseline data and fully adjusted models that include relevant facility readiness indices, sociodemographic characteristics and province fixed effects, as described in the second regression model in Section 4.2.

Good perception of quality of care was associated with higher ANC1, less late initiation of ANC, high PNC, and increased likelihood of discussion complications during ANC visits. It was not associated with ANC4 or likelihood to experience complications. Women with a better facility nearby was associated with higher ANC1 and PNC rates, and were more likely to discussion complications. It was not associated with timely initiation of ANC, completion of adequate ANC, or likelihood to experience complications.

**Any antenatal care (ANC1):**

- Women with a “good” perception of care quality were 44% more likely to attend at least one ANC visit (OR = 1.44, 95% CI: 1.13–1.84).
- Women attending facilities in the top-performing tier for prenatal services were also more likely to seek ANC (OR = 1.21, 95% CI: 1.01–1.45).

**Late initiation of ANC:**

- Better perceived quality was associated with a 20% reduction in the odds of delaying the first visit (OR = 0.80, 95% CI: 0.66–0.98).
- No facility readiness domain was significantly associated with this outcome.

**Adequate ANC (4+ visits):**

- Perceived quality was not statistically associated with this outcome (OR = 0.88, 95% CI: 0.72–1.06).
- No facility readiness indicators reached significance for this outcome.

**Any postnatal care (PNC):**

- Women with high perceived quality had 70% higher odds of receiving postnatal care (OR = 1.70, 95% CI: 1.20–2.40).
- Use was also higher among women attending top-performing facilities for prenatal services (OR = 1.34, 95% CI: 1.06–1.68) and malaria services (OR = 1.41, 95% CI: 1.09–1.83).

**Discussion of complications during ANC:**

- Women with positive perceptions were over twice as likely to report discussing complications (OR = 2.43, 95% CI: 1.91–3.11).
- Women attending facilities with strong performance in childbirth and postpartum services were also more likely to engage in these discussions (OR = 1.26, 95% CI: 1.05– 1.51).

**Experienced complications during pregnancy or delivery:**

- Neither perceived quality nor facility readiness indicators were significantly associated with this outcome (see Appendix C for additional results on food insecurity and patient experience).

Results from the endline household survey showed similar associations as those observed at baseline. At endline, women with a good perception of care were 56% more likely to attend at least one ANC visit (OR = 1.56, 95% CI: 1.09–2.22), 23% more likely to complete four or more ANC visits (OR = 1.23, 95% CI: 1.01–1.51), 30% less likely to delay the start of antenatal care (OR = 0.70, 95% CI: 0.57–0.87), and 38% less likely to initiate postnatal care late (OR = 0.62, 95% CI: 0.39–0.98).

When disaggregating the endline analysis by financing model, associations between perceived quality of care and health-seeking behavior remained strong and statistically significant in DFF settings (late ANC initiation, late PNC initiation, PNC use, and discussion of complications; see Table 2 for reference). In contrast, associations were weaker and mostly insignificant in PBF settings, with the exception of ANC1 attendance (see Table 2 for reference).

**Table 2.**
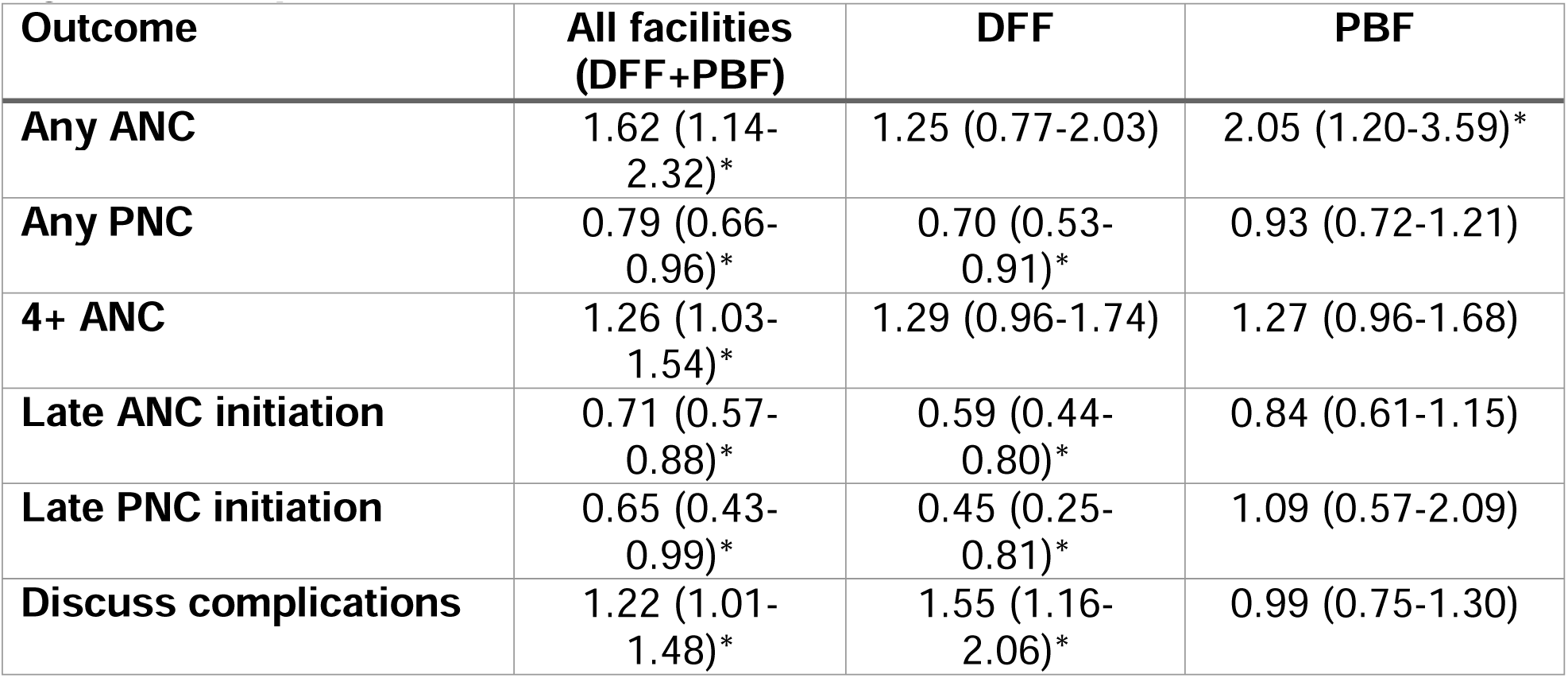
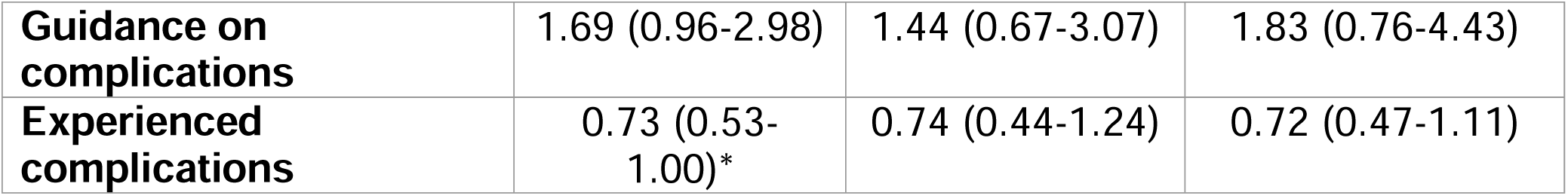
Adjusted Odds Ratios and 95% Confidence Intervals for the Association Between Perceived Quality of Care and Maternal Health Outcomes, by Facility Financing Model. *Odds of a pregnant person with a “good” overall perception of care having an outcome, compared to someone with a “poor” perception. All models adjust for relevant facility readiness domains and individual-level covariates. DFF = direct facility financing. PBF = performance-based financing. ANC = antenatal care. PNC = postnatal care. Late ANC = initiated after six months pregnant. Late PNC = initiated after seven days postpartum. *Indicates statistical significance at the p < 0.05 level*.

### Effects of Financing Model on Facility Behavior and Quality Improvement

We next examine how the type of financing model influenced facility performance over time, focusing on relative improvements between baseline and endline as described in section 4.3. By comparing changes in service readiness and perceived quality across performance-based financing (PBF) and direct facility financing (DFF), we assess how conditional versus unconditional funding shaped facility performance.

Figure 4 shows the relative percentage change in key domains, with PBF facilities experiencing significantly greater improvements in vaccination and cold chain services (+56.7%, 95% CI: 39.9% to 73.5%), laboratory test availability (+35.1%, 95% CI: 15.4% to 54.7%), tuberculosis services (+48.2%, 95% CI: 21.9% to 74.6%), and prenatal services (+19.2%, 95% CI: 8.3% to 30.2%) compared to DFF facilities. However, malaria services improved 25.2% more under DFF than under PBF (95% CI: −38.9% to −11.4%).

**Figure 4.**
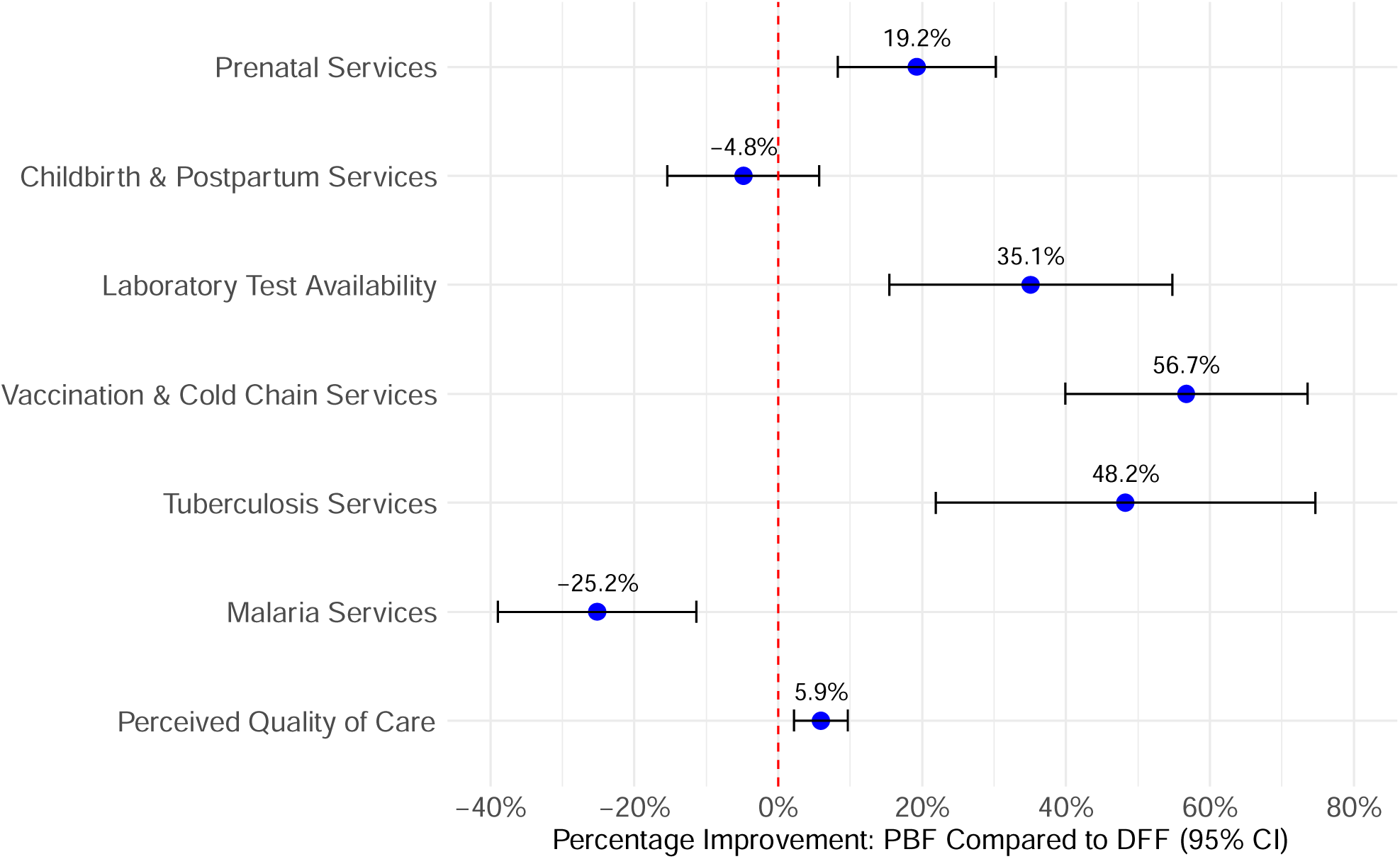
Relative Improvements in Service Readiness and Perceived Quality Under PBF Compared to DFF. *Normalized Difference-in-differences estimates of changes from baseline to endline among facilities observed in both rounds. Positive values indicate greater improvement under performance-based financing (PBF); negative values indicate greater improvement under direct facility financing (DFF). Error bars represent 95% confidence intervals. CI = confidence interval*.

We also observe a modest but significant improvement in average perceived quality of care— aggregated at the facility level from household responses—under PBF relative to DFF (+5.9%, 95% CI: 2.2% to 9.7%).

### Financial Hardship, Perceived Affordability, and Antenatal Care Use

To quantify how financial hardship constrains maternal care-seeking, we decomposed its impact into direct and indirect pathways through a mediation framework. This analysis builds on the approach outlined in Section 4.4 and focuses on whether hardship indicators reduce antenatal care attendance directly or by first shaping affordability perceptions in the baseline household survey.

When women perceive service costs as acceptable, they are 47% more likely to attend at least one antenatal care (ANC) visit. However, among women whose health expenses exceeded their income, the effect of perceived affordability was attenuated by 19.7 percentage points, meaning that their likelihood of seeking care dropped to 28%. In this case, the total effect of financial hardship on ANC1 was 28%, driven entirely by the indirect pathway through perceived service cost acceptability. There was no statistically significant direct effect, indicating that hardship influences care-seeking primarily by shaping perceptions (see Table 3 in the Supplemental Material for results using service cost acceptability; includes estimates for two additional financial hardship metrics: receiving a gift and borrowing for health expenses). A similar pattern holds for medicine cost acceptability. Initially, women who found medicine costs acceptable were 43% more likely to seek ANC. However, financial hardship again lowered this perception, reducing the effect by 15.6 percentage points, resulting in a total effect of 28% (see Table 4 in the Supplemental Material for results using medicine cost acceptability; includes estimates for two additional financial hardship metrics: receiving a gift and borrowing for health expenses).

Finally, to complement this analysis, we also explored how households respond to financial strain through coping mechanisms-recognizing that households facing high health expenditures (i.e., health expenditures that exceeded income) may resort to selling food reserves, livestock, or agricultural equipment as a coping mechanism. Selling food reserves was significantly associated with a decreased likelihood of late ANC initiation (OR = 0.45, 95% CI: 0.25–0.82), suggesting that this strategy may enable early care-seeking in the face of financial hardship.. However, it was also linked to reduced odds of receiving adequate antenatal care -ANC4-(OR = 0.44, 95% CI: 0.26–0.76) and lower likelihood of postnatal care (OR = 0.36, 95% CI: 0.14–0.91), indicating that the short-term benefits of this coping mechanism may not enable care continuity. In contrast, selling agricultural equipment was associated with increased odds of receiving adequate antenatal care -ANC4-(OR = 1.85, 95% CI: 1.23–2.79) and postnatal care (OR = 1.89, 95% CI: 1.08–3.31), without any significant effect on late ANC initiation or ANC1. This suggests that while selling agricultural equipment may not influence the timing of care initiation, it may be more effective at sustaining maternal healthcare engagement across later stages such as adequate ANC and postnatal care. No statistically significant associations were observed between livestock sales and any of the maternal health outcomes (see Figure 5 for reference).

**Figure 5.**
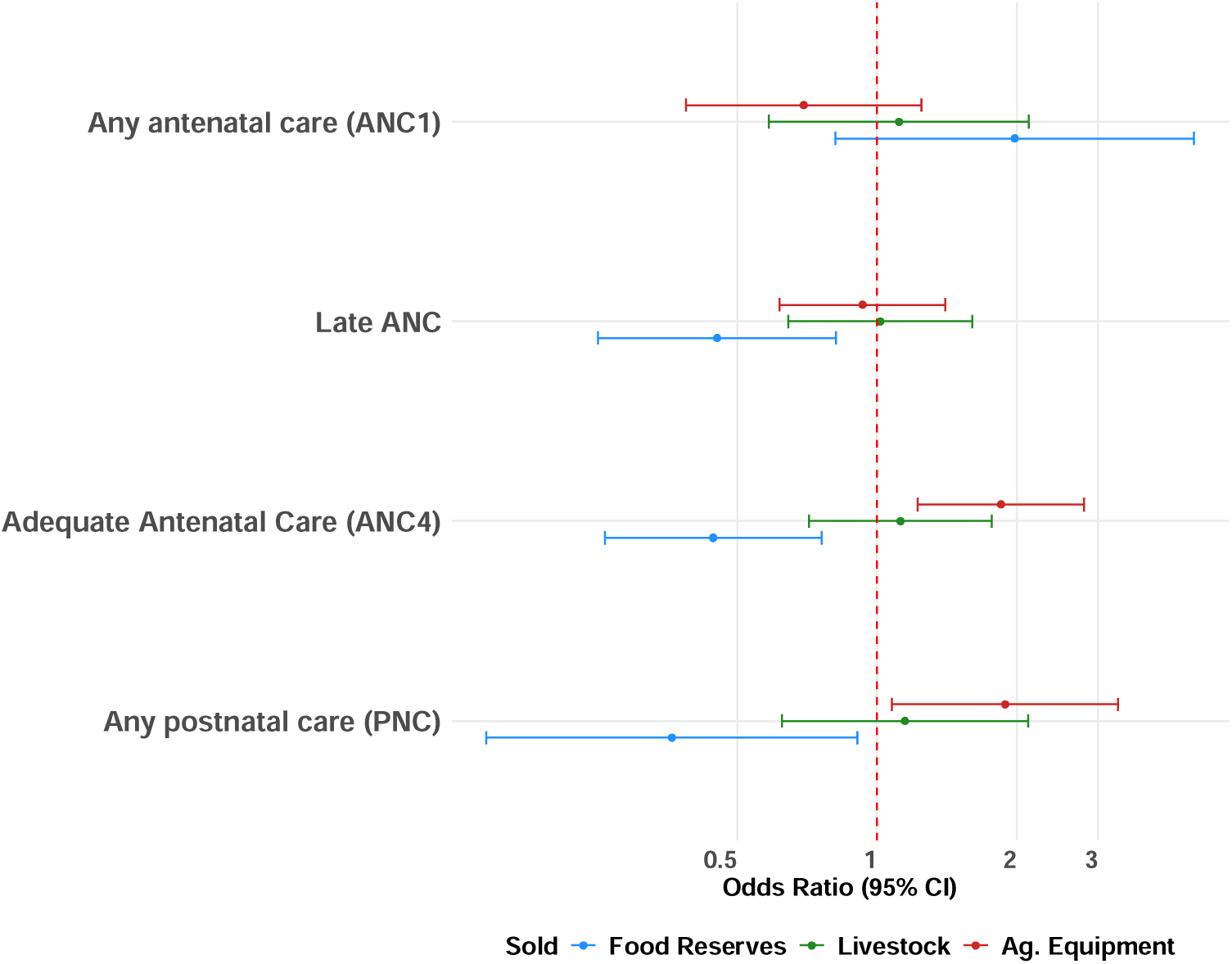
Association Between Types of Asset Sales and Maternal Health-Seeking Behaviors. *Odds ratios and 95% confidence intervals for selling food reserves, livestock, or agricultural equipment on four outcomes: any ANC, late ANC, adequate ANC (ANC4), and any postnatal care (PNC). CI = confidence interval*.

## Discussion

Our analysis reveals that maternal health-seeking behavior in the DRC is driven less by physical infrastructure or observable facility readiness, and more by how women perceive and experience care within the health system. Across two rounds of linked household and facility data, perceived quality of care consistently emerged as the strongest predictor of key outcomes—including initiation of ANC, receipt of PNC, and discussing potential complications. Women who reported respectful treatment, clear communication, and cleanliness and courtesy were significantly more likely to initiate care with the health system, independent of objective facility readiness. Overall, for ANC1, positive perceptions increased the odds of care seeking by +29-61% (odds ratio) and were significant for 85% of the perceptions that were evaluated. For PNC, positive perceptions increased the odds of care seeking by +63-316% (odds ratio) and were significant for 77% of the perceptions that were evaluated.

Using the same dataset, others have found that supply-side readiness has modest impact on ANC clinical quality. [18] However, this was limited to only looking at women who had already chosen to attend ANC, and our findings here suggest that structural readiness alone is insufficient to drive care-seeking without corresponding improvements in patient experience and perceived responsiveness. This highlights the individual-dependent nature of health-seeking behavior, where outcomes hinge on interpersonal trust and perceived responsiveness of care.

Similarly, perceptions of affordability were more predictive of care utilization than direct measures of financial hardship—a dynamic shaped by both individual-level expectations and population-level economic vulnerability. This relationship was further reflected in the specifics of coping behaviors: women in financially constrained households often sold assets to pay for care. The type of asset sold—such as food reserves versus agricultural equipment—was associated with distinct care trajectories, suggesting that the economic decisions available to a household (i.e., the kind of assets they own) affect their choices, and how they make trade-offs depends on the anticipated benefit of getting medical care. Taken together, the results indicate that individuals’ perceptions of healthcare services—more than structural readiness or explicit costs—are central determinants of maternal care-seeking behavior. Understanding how these perceptions are formed, reinforced, and transmitted is therefore critical—not only because they guide personal decisions, but because they shape collective trust and community norms around service use.

These perceptions are not secondary or peripheral aspects of health systems—they are fundamental drivers of care utilization. Women’s perceptions of quality of care are formed primarily through tangible interpersonal experiences with providers (e.g., being treated with respect, receiving clear explanations, feeling heard) and through direct assessments of facility conditions (e.g., cleanliness, drug availability, and confidence in the system’s ability to deliver effective care). In fragile health systems, these impressions carry disproportionate weight. If care is perceived as unsafe, disrespectful, or futile, women may forgo it—even when services are technically available and helpful. Conversely, when care is perceived as responsive, kind, and trustworthy, it can catalyze care-seeking and continued engagement.

This dynamic becomes even clearer when recognizing that no interaction with the health system exists in isolation: a negative experience during antenatal care can undermine trust in subsequent skilled deliveries or childhood immunizations, and one woman’s encounter with disrespect or stock-outs can ripple through her social network, discouraging others from seeking care.

Moreover, in places with pervasive financial insecurity such as rural DRC, these perceptions are further shaped by households’ ongoing evaluations of whether accessing care is justified, given their limited resources.

As expected, perceived unaffordability reduces care-seeking. More telling, however, are the specific coping strategies households employ—such as selling food reserves or livestock—which reveal both economic vulnerability and varying levels of commitment to pursuing care. Our findings reflect this nuanced behavioral calculus, echoing broader calls for a “quality revolution” that positions trust, person-centeredness, and safety not as optional extras, but as essential pillars of effective health systems [19], [20], [21] that can both improve the actual care delivered and increase coverage. Consistent with this vision, our results underline the importance of improving perceptions of quality, which requires both investments in the experience of care (e.g., improved cleanliness) as well as communicating improvements that have occurred (e.g., more convenient open hours). This will ensure that health systems are not merely accessed, but trusted, valued, and sustainably utilized.

Beyond these individual and perception-based determinants, our findings highlight how health financing structures also shape maternal care-seeking behavior. Specifically, we observe notable differences in women whose closest facility was either receiving PBF or DFF, suggesting that the chosen financing model influences not only the types of services delivered but also how these services are experienced by patients and perceived by communities.

In DFF facilities, women’s perceptions play a more significant role in their decisions to seek care. Without strict performance-based conditions, facility responsiveness to improving patient experience may vary considerably, thereby elevating the importance of patient perception as an indicator of quality. As a result, women’s subjective experiences and assessments become central factors guiding their decisions about whether to initiate or continue care. By contrast, PBF models standardize service provision around specific incentivized indicators, reducing variability in service delivery across facilities. This standardization can diminish the relative influence of patient perceptions in shaping care-seeking behavior. Thus, financing models not only shape how providers deliver services but also influence how patients perceive and respond to care.

Furthermore, we observed that DFF facilities often performed particularly well in service areas aligned with high-burden local priorities, such as malaria care. Compared to PBF, where centrally defined incentive structures guide investments, the greater flexibility in resource allocation under DFF may encourage facilities to respond more directly to local epidemiological conditions and community-specific health needs. This complements the more standardized improvements observed under PBF—particularly in vaccination, tuberculosis, prenatal and postnatal services—where performance indicators were more explicitly targeted [22].

These findings align with the recommendations of the 2022 Lancet Commission on Primary Health Care Financing, which emphasized that local autonomy, flexibility, and facility-level accountability are critical components of high-quality primary care systems [23]. According to the Commission, overly rigid financing frameworks may limit a facility’s ability to address localized health priorities, especially where the epidemiology and social barriers are variable and require adaptive responses. More broadly, this contrast between DFF and PBF shows how financing models shape not only how well the health system performs, but also how patients experience and respond to care being made available. Given that expressed demand is not equivalent to local health needs, this complexity needs to be further explored and reflects a systems-dependent dynamic, where the structure and design of the delivery platform play a key role in determining the success of an intervention.

We note that while these findings offer important insights into how perceptions and financing structures shape maternal care-seeking behavior and reinforce previous results, they should be interpreted with caution. The use of cross-sectional data at each time point limits causal inference and prevents us from observing within-person changes over time. Temporal ambiguity in key survey items—such as the sequencing of complications and provider interactions—further constrains our ability to determine directionality. Additionally, differences in geographic coverage between baseline and endline may introduce unrelated variation, limiting comparability across timepoints.

Our findings demand a redefinition of health system performance, placing patient experience firmly alongside structural readiness and clinical quality as a core objective—not merely as an indicator. Respectful, responsive, and trusted care should be explicitly tracked as critical components of high-quality service delivery. Metrics capturing trust, interpersonal quality, and perceptions of affordability must be integrated systematically into routine quality assessments, using independently collected data to ensure objectivity and prevent bias. Additionally, tracking the types of assets households sell and the proportion experiencing financial hardship could provide actionable insights into perceived affordability and economic vulnerabilities that influence care-seeking behaviors.

Financing models could incentivize these patient-centered metrics, but caution is required to avoid perverse incentives, such as providers pressuring patients into favorable reporting—a practice that can rapidly undermine the trust that we have found to be so important. Crucially, efforts to scale DFF should integrate incentives explicitly tied to facility responsiveness to patient feedback, reinforcing frontline autonomy and adaptability. Positioning DFF as a genuinely people-centered delivery platform contrasts favorably with more rigid PBF approaches, making health systems not only more responsive, but fundamentally more trusted by the communities they serve.

Contrary to traditional approaches to investing in primary healthcare, substantially greater emphasis must be placed on factors shaping patient perceptions—friendliness, cleanliness, and affordability—if we aim to fundamentally improve coverage. Perception data is not merely a satisfaction score; it is a proxy for trust, a powerful predictor of utilization, and the critical missing link in primary health care systems that aspire to be genuinely people centered.

## Supplemental Results

**Table 3.**
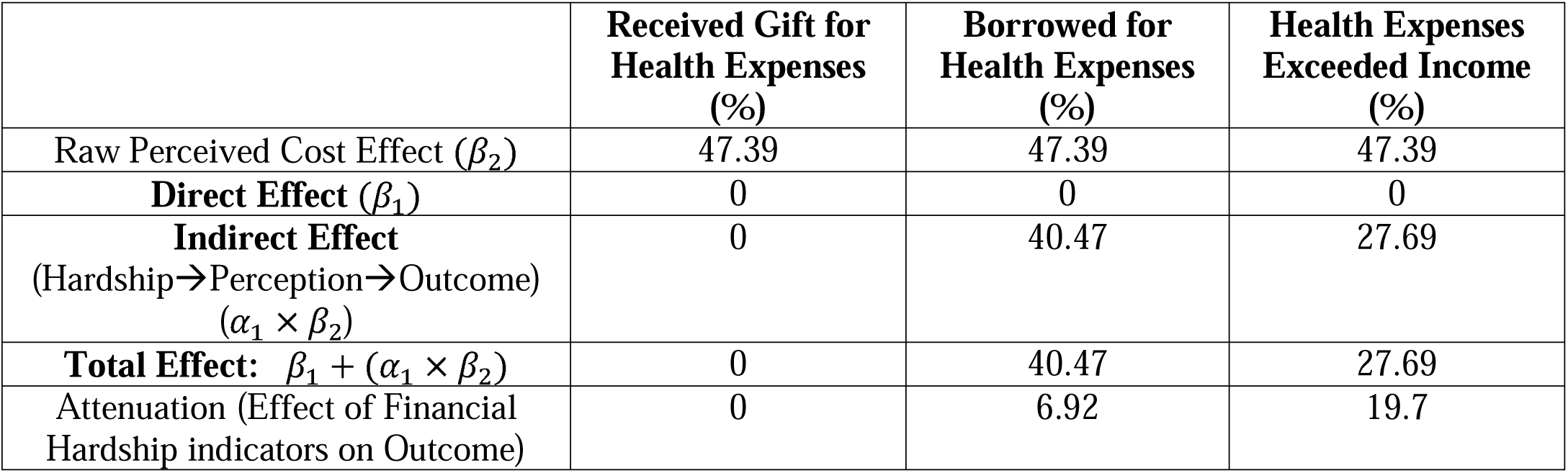
Decomposition of the Effect of Financial Hardship on Antenatal Care Use via Service Cost Acceptability. *Values represent the direct, indirect, and total effects of financial hardship indicators on the likelihood of seeking any antenatal care (ANC1), with service cost acceptability as the perceived cost variable. The indirect effect captures the pathway through perceived affordability. All effects are reported as percentage point changes*.

|  | Received Gift for<br>Health Expenses<br>(%) | Borrowed for<br>Health Expenses<br>(%) | Health Expenses<br>Exceeded Income<br>(%) |
| --- | --- | --- | --- |
| Raw Perceived Cost Effect ($\beta_2$) | 47.39 | 47.39 | 47.39 |
| **Direct Effect** ($\beta_1$) | 0 | 0 | 0 |
| **Indirect Effect**<br>(Hardship→Perception→Outcome)<br>($\alpha_1 \times \beta_2$) | 0 | 40.47 | 27.69 |
| **Total Effect:** $\beta_1 + (\alpha_1 \times \beta_2)$ | 0 | 40.47 | 27.69 |
| Attenuation (Effect of Financial<br>Hardship indicators on Outcome) | 0 | 6.92 | 19.7 |

**Table 4.** Decomposition of the Effect of Financial Hardship on Antenatal Care Use via Medicine Cost Acceptability. *Values represent the direct, indirect, and total effects of financial hardship indicators on the likelihood of seeking any antenatal care (ANC1), with medicine cost acceptability as the perceived cost variable. The indirect effect captures the pathway through perceived affordability. All effects are reported as percentage point changes*.

|  | Received Gift for<br>Health Expenses<br>(%) | Borrowed for<br>Health Expenses<br>(%) | Health Expenses<br>Exceeded Income<br>(%) |
| --- | --- | --- | --- |
| Raw Perceived Cost Effect ($\beta_2$) | 43.27 | 43.27 | 43.27 |
| **Direct Effect** ($\beta_1$) | 0 | 0 | 0 |
| **Indirect Effect**<br>(Hardship→Perception→Outcome)<br>($\alpha_1 \times \beta_2$) | 0 | 36.88 | 27.65 |
| **Total Effect:** $\beta_1 + (\alpha_1 \times \beta_2)$ | 0 | 36.88 | 27.65 |
| Attenuation (Effect of Financial<br>Hardship indicators on Outcome) | 0 | 6.39 | 15.62 |

## Acknowledgements

The authors would like to thank Gil Shapira and Supriya Madhavan for their time and insight into the context and implementation details of the PDSS program.

## Data Availability

No data was generated for this study. All data used for analysis is available at the World Bank microdata library, under codes COD_2015_HRBFIE-FBL_v01_M, COD_2015_HRBFIE-HBL_v01_M, COD_2021-2022_HRBFIE-HFFU_v01_M, and COD_2021-2022_HRBFIE-HHFU_v01_M.

## Appendix A. Validation of Perceived Quality of Care Index

To ensure that the index appropriately reflects patterns in the underlying perception items, we conducted a validation exercise using the baseline household survey data. Figure A1 displays the average values of each perception item across the three index-defined categories. As expected, women classified as reporting “good” perceived quality had consistently higher scores across all individual perception items, while those classified as “poor” had the lowest values. This alignment supports the internal consistency and construct validity of the index.

**Figure A1.**
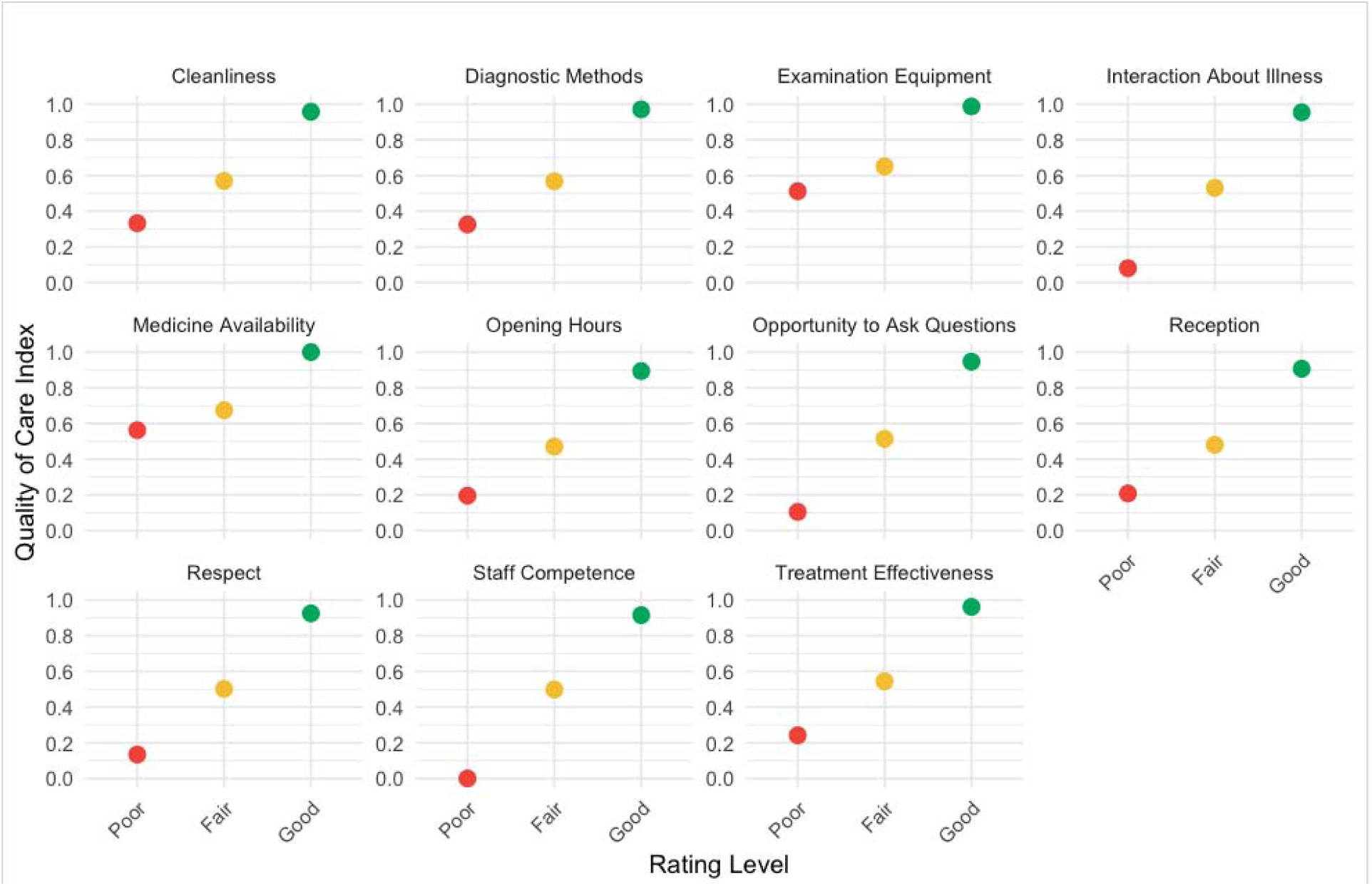
Validation of Perceived Quality of Care Index Using Underlying Survey Items.

## Appendix B. Overview of Facility Readiness Indices

| Domain | Summary of Information Included |
| --- | --- |
| **Laboratory Test Availability** | Availability of diagnostic tests (e.g., HIV, syphilis, hemoglobin, malaria RDT), lab equipment (e.g., microscopes, centrifuges), operating hours, and presence of lab staff and registers |
| **Vaccination & Cold Chain Services** | Provision of vaccination services, cold chain equipment (fridges, carriers), temperature monitoring practices, service hours, and community outreach activities |
| **Prenatal Services** | Availability of ANC services, weekday and weekend hours, provision of supplements and preventive medications (e.g., iron, folic acid, SP), and counseling services |
| **Childbirth & Postpartum Services** | Availability of Emergency Obstetric and Newborn Care (EmONC) signal functions, obstetric medications (e.g., oxytocin, anticonvulsants), postpartum counseling, and frequency of postnatal consultations |
| **Tuberculosis Services** | TB service availability, infrastructure (dedicated room), availability of treatment registers, DOTS strategy implementation, and staff training |
| **Malaria Services** | Case detection and confirmation practices (RDT and lab), treatment coverage with ACTs, ITN availability, and adherence to national guidelines |
| **Essential Medicines Availability** | Availability of a wide range of tracer essential medicines (e.g., antibiotics, antimalarials, analgesics, iron, ORS, vitamin A) and basic consumables (e.g., gloves, syringes) |
| **Operational Equipment** | Presence and functionality of essential clinical tools (e.g., blood pressure monitors, weighing scales, stethoscopes, oxygen supply, resuscitation equipment, surgical kits, IV catheters) |

## Appendix C. Food Insecurity, Iron Supplementation, and Pregnancy Outcomes

Food insecurity was associated with more active health-seeking behavior and engagement during antenatal care. Women facing food insecurity were significantly more likely to discuss complications during ANC visits (OR = 1.72, 95% CI: 1.29–2.29) and to take proactive steps to address illness (OR = 1.40, 95% CI: 1.07–1.83), compared to food-secure women. However, despite this increased awareness and behavioral responsiveness, food-insecure women remained more likely to report complications during pregnancy or delivery (OR = 1.36, 95% CI: 1.05– 1.77). This suggests that awareness and proactive measures alone may not be sufficient to mitigate underlying risks.

Among food-secure women, taking iron tablets reduces complications (coefficient: –0.058, *p* = 0.06). However, iron tablets cannot overcome the general malnutrition of food-insecure women, resulting in no net effect of iron tablets on complications in this population. In other words, the elevated baseline risk of complications among food-insecure women cancels out the protective effect of iron tablets, resulting in no net improvement in outcomes (interaction term of food insecurity and iron tables, +0.068, *p* = 0.04).

Importantly, both food-secure and food-insecure women reported similar durations of iron use during pregnancy-16 days (95% CI: 3–30) versus 18 days (95% CI: 2–60), respectively (*p* < 0.001)-indicating that differences in reported intake are unlikely to explain the gap in outcomes. These findings suggest that iron alone may be insufficient for reducing complications among nutritionally vulnerable women, and that more comprehensive interventions-such as multiple micronutrient supplementation-may be needed to address these disparities.

## References

[1] W. H. Organization and others, “Trends in maternal mortality 2000 to 2017: estimates by WHO, UNICEF, UNFPA, World Bank Group and the United Nations Population Division: executive summary,” 2019.

[2] W. Graham et al., “Diversity and divergence: the dynamic burden of poor maternal health,” The Lancet, vol. 388, no. 10056, pp. 2164–2175, 2016.

[3] M. du P. et Suivi, “de la Mise en œuvre de la Révolution de la Modernité (MPSMRM), Ministere de la Santé Publique (MSP) and ICF International. 2014,” Enquête Démographique et de Santé en République Démocratique du Congo, vol. 2014, 2013.

[4] INS, “Enquête par grappes à indicateurs multiples, 2017–2018, rapport de résultats de l’enquête,” 2018, Kinshasa, République Démocratique du Congo.

[5] D. R. Hotchkiss, L. S. Blum, L. S. Craig, A. Yemweni, J. Wisniewski, and P.-S. Lusamba-Dikassa, “Assessing the impact of complex health systems strengthening programs on maternal health care utilization in fragile and conflict-affected states: evidence from the Democratic Republic of the Congo,” BMC Pregnancy Childbirth, vol. 25, no. 1, pp. 1–16, 2025.

[6] M. Koblinsky et al., “Quality maternity care for every woman, everywhere: a call to action,” The Lancet, vol. 388, no. 10057, pp. 2307–2320, 2016.

[7] B. R. Ziegler, M. Kansanga, Y. Sano, J. Kangmennaang, D. Kpienbaareh, and I. Luginaah, “Antenatal care utilization in the fragile and conflict-affected context of the Democratic Republic of the Congo,” Soc Sci Med, vol. 262, p. 113253, 2020.

[8] D. Bwirire, I. Roosen, N. De Vries, R. Letschert, E. Ntabe Namegabe, and R. Crutzen, “Maternal Health Care Service Utilization in the Post-Conflict Democratic Republic of Congo: An Analysis of Health Inequalities over Time,” in Healthcare, 2023, p. 2871.

[9] Y. Akachi and M. E. Kruk, “Quality of care: measuring a neglected driver of improved health,” Bull World Health Organ, vol. 95, no. 6, p. 465, 2016.

[10] S. Roder-DeWan et al., “Expectations of healthcare quality: a cross-sectional study of internet users in 12 low-and middle-income countries,” PLoS Med, vol. 16, no. 8, p. e1002879, 2019.

[11] G. Thapa, M. Jhalani, S. Garc\’\ia-Saisó, A. Malata, S. Roder-DeWan, and H. H. Leslie, “High quality health systems in the SDG era: country-specific priorities for improving quality of care,” PLoS Med, vol. 16, no. 10, p. e1002946, 2019.

[12] Y. Akachi and M. E. Kruk, “Quality of care: measuring a neglected driver of improved health,” Bull World Health Organ, vol. 95, no. 6, p. 465, 2016.

[13] A. Austin, A. Langer, R. A. Salam, Z. S. Lassi, J. K. Das, and Z. A. Bhutta, “Approaches to improve the quality of maternal and newborn health care: an overview of the evidence,” Reprod Health, vol. 11, pp. 1–9, 2014.

[14] S. Bhattacharyya, A. Srivastava, M. Saxena, M. Gogoi, P. Dwivedi, and K. Giessler, “Do women’s perspectives of quality of care during childbirth match with those of providers? A qualitative study in Uttar Pradesh, India,” Glob Health Action, vol. 11, no. 1, p. 1527971, 2018.

[15] S. K. Acharya, S. K. Sharma, B. P. Dulal, and K. K. Aryal, “Quality of care and client satisfaction with maternal health services in Nepal,” Rockville, Maryland, USA, 2018. [Online]. Available: http://dhsprogram.com/pubs/pdf/FA112/FA112.pdf

[16] J. P. Souza et al., “Moving beyond essential interventions for reduction of maternal mortality (the WHO Multicountry Survey on Maternal and Newborn Health): a cross-sectional study,” The Lancet, vol. 381, no. 9879, pp. 1747–1755, 2013.

[17] A. Yoseph, W. Teklesilasie, F. Guillen-Grima, and A. Astatkie, “Perceptions, barriers, and facilitators of maternal health service utilization in southern Ethiopia: A qualitative exploration of community members’ and health care providers’ views,” PLoS One, vol. 19, no. 12, p. e0312484, 2024.

[18] R. Han and B. Hagedorn, “More than the sum of its parts? An analysis of the factors impacting the content of antenatal care consultations in the Democratic Republic of Congo,” *medRxiv*, pp. 2021–2025, 2025.

[19] M. E. Kruk, E. Larson, and N. A. Y. Twum-Danso, “Time for a quality revolution in global health,” Lancet Glob Health, vol. 4, no. 9, pp. e594–e596, 2016.

[20] M. E. Kruk, M. Pate, and Z. Mullan, “Introducing the Lancet Global Health Commission on high-quality health systems in the SDG era,” Lancet Glob Health, vol. 5, no. 5, pp. e480–e481, 2017.

[21] M. E. Kruk and M. Pate, “The lancet global health commission on high quality health systems 1 year on: progress on a global imperative,” Lancet Glob Health, vol. 8, no. 1, pp. e30–e32, 2020.

[22] G. Shapira et al., “Impacts of performance-based financing on health system performance: evidence from the Democratic Republic of Congo,” BMC Med, vol. 21, no. 1, p. 381, 2023.

[23] K. Hanson et al., “The Lancet Global Health Commission on financing primary health care: putting people at the centre,” Lancet Glob Health, vol. 10, no. 5, pp. e715–e772, 2022.

